# Portable nucleic acid tests for rapid detection of monkeypox virus

**DOI:** 10.1101/2022.08.09.22278605

**Authors:** Sanchita Bhadra, Andrew D. Ellington

**Author notes:** Emails: SB; ADE.

## Abstract

We report two isothermal nucleic acid amplification assays for detection of monkeypox virus (MPXV) clades 2 and 3 that include the strains responsible for the current global outbreak of monkeypox. The assays use loop-mediated isothermal amplification (LAMP) to amplify two distinct sequences in the MPXV genome. Readout specificity is ensured by oligonucleotide strand displacement (OSD) probes integrated in one-pot LAMP-OSD reactions. OSD probes undergo toehold-mediated strand displacement hybridization to LAMP amplicon loop sequences derived from MPXV clades 2 and 3 resulting in fluorescence readable both in realtime and visually at endpoint. We also perform both assays on two different portable devices, the GeneTiger and the miniPCR, to exemplify compatibility with minimum infrastructure point-of-care (POC) testing in clinical and at-home settings. Both assays could readily detect single digit copies of MPXV synthetic double stranded DNA templates within 30 min.

---

Monkeypox is a zoonotic smallpox-like disease characterized by fever and rashes on skin and mucosal membranes [1]. It is caused by the Monkeypox virus (MPXV), a large enveloped double stranded DNA virus belonging to the genus *Orthopoxvirus* in the *Poxviridae* family [1]. MPXV is currently classified into three clades – the more virulent and transmissible clade 1 (formerly termed the Central African or Congo Basin clade), clade 2 (formerly the West African clade), and clade 3 that includes most genomes from the 2017 - 2022 human outbreaks (formerly also in the West African clade) [2, 3]. Animal-to-human transmission usually occurs by direct contact with contaminated body fluids or lesions while human-to-human transmission can also occur via close contact with infected skin lesions, objects, and respiratory secretions. Many animals, including non-human primates, are susceptible to MPXV however, the natural reservoir(s) and the circulation cycle of the virus remain undetermined [1]. Historically, most cases were reported from rural regions of central and west Africa. Since the first monkeypox outbreak reported outside Africa in 2003, MPXV has increasingly become a global health concern. As of writing this manuscript, 30,189 cases of monkeypox have been confirmed in 88 countries of which 29,844 cases were in 81 countries with no previous reports of the disease [4].

Expeditious and accurate identification of new cases is crucial for preventing and reducing viral spread. Moreover, due to the frequent attachment of stigma to monkeypox diagnosis, access to reliable self-testing methods would enable a wider range of public health options [5]. Since Orthopoxviruses are serologically cross-reactive and antigen/antibody-based tests can generate false positive outcomes, polymerase chain reaction (PCR) assays are the diagnostic standard. PCR-based tests are usually implemented on complex instruments within centralized clinical laboratory infrastructures that require sample shipment and several hours to weeks of sample-to-result turnaround time. Isothermal nucleic acid amplification tests (iNAT), such as loop-mediated isothermal amplification (LAMP), that do not need thermocyclers and therefore can be simpler to operate at point-of-care (POC), have been reported for MPXV [6]. However, the use of increasing reaction turbidity due to Mg^2+^ precipitation as the indicator of amplicon accumulation renders this assay susceptible to false positive readout of spurious amplification, a relatively common caveat of isothermal amplification techniques. Other isothermal tests with nucleic acid probe-based readout of recombinase polymerase amplification (RPA) products have also been developed for MPXV [7] however, RPA can be more expensive than LAMP due to its dependence on commercial multi-protein reaction mixes and probe oligonucleotides with multiple chemical modifications. In contrast, LAMP reaction mixes can be readily assembled using customizable homemade buffers and enzymes. Bst DNA polymerase large fragment, the workhorse strand displacing DNA polymerase used for LAMP, can be sourced commercially or readily purified in-house using publicly available protein expression constructs and purification protocols (Addgene Plasmid #145799). In fact, we have shown that instead of purified enzymes LAMP-based diagnostics can be performed using ‘cellular’ reagents comprising dried bacteria overexpressing Bst-LF [8-10]. Moreover, we have recently developed and made available a suite of engineered Bst-LF derived enzymes with improved temperature and chaotrope tolerance and enhanced reverse transcription abilities that can be used to configure faster single enzyme sample-to-answer LAMP detection of DNA and RNA targets ([11, 12]; Addgene Plasmids # 161875, 179278, 171200, and 171201).

Along with the strand displacing DNA polymerase, LAMP typically uses at least two inner primers, termed FIP and BIP, and two outer primers, termed F3 and B3, specific to 6 different regions (F3, F2, F1, B1, B2, and B3) of the target DNA or RNA, to produce 10^9^ to 10^10^ concatemeric amplicons within one hour [13]. This rapid amplification is facilitated by the ability of FIP and BIP sequences at the 3’- and 5’- ends of LAMP amplicons to foldback into stem loop structures that bind new inner primers and enable self-priming of strand displacement amplification. Additional loop-specific primers (LP), stem primers, and swarm primers can be used to accelerate amplification [14-16]. This continuous and exponential amplification process allows LAMP to often rival the detection limit of PCR, however non-specific primer interactions can also be rapidly amplified yielding spurious amplicons that cause non-specific readout methods, such as Mg^2+^ precipitation [17], fluorescent dye intercalation [18], and pH changes [19], to produce false positive outcomes. To improve LAMP readout accuracy, we previously used concepts from the field of DNA computation [20] to develop oligonucleotide strand displacement (OSD) probes that, similar to TaqMan probes in PCR [21], produce a signal only in the presence of correct LAMP amplicons [22]. OSDs are complementary to one of the loop sequences of the correct LAMP amplicon and do not overlap primer binding sites. They are comprised of a long strand and a complementary short strand that both lack 3’-OH groups and cannot serve as primers. In the absence of the correct LAMP amplicons, the two OSD strands form a hemiduplex in which a short single stranded region, termed toehold, remains exposed at either the 5’- or the 3’-end of the long strand (**Figure 1**). When a correct LAMP amplicon is available, this toehold hybridizes to its complementary LAMP loop and initiates branch migration that causes the long strand to progressively separate from the short strand and instead hybridize to the LAMP loop (**Figure 1**). This strand displacement event, indicative of the correct LAMP amplicon, can be measured using a variety of readout modes, such as fluorimetry and colorimetric lateral flow assays [23, 24].

**Figure 1.**
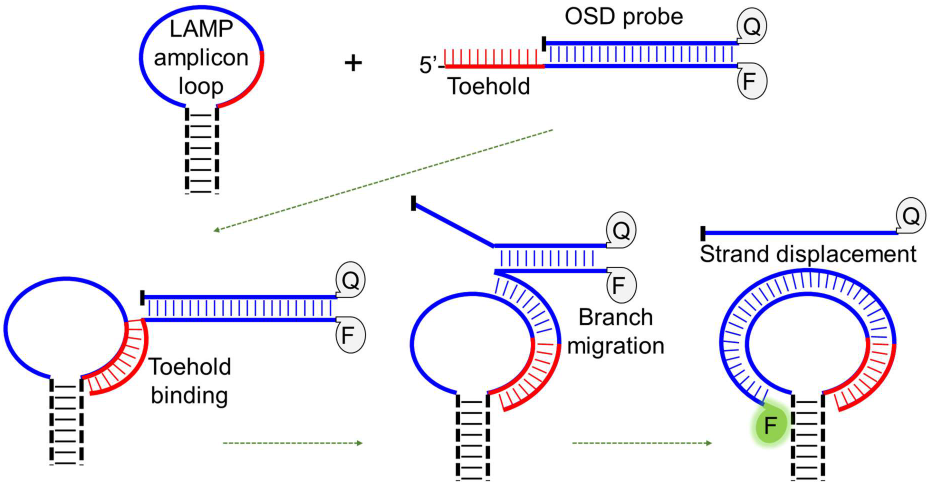
Schematic depicting toehold-mediated strand displacement hybridization of an OSD probe and a LAMP amplicon loop. The bar at the 3’-end of the OSD short strand represents the blocked 3’-OH group that prevents polymerase-mediated extension. ‘F’: covalently attached fluorophore; ‘Q’: covalently attached quencher moiety.

With over 20 published assays, compatibility with one-pot direct analysis of crude samples, such as environmental water, crushed insects, and human saliva, capacity for template semi-quantitation, and ability to logically process amplicon signal, such as distinguishing single nucleotide polymorphisms and calculating co-presence of multiplex amplicons, LAMP-OSD has proven to be a robust and versatile assay platform [22-29]. We now report two clades 2 and 3 MPXV LAMP-OSD assays for rapid diagnosis of the ongoing monkeypox outbreak [3]. Furthermore, we demonstrate both assays on two different portable diagnostic devices to exemplify suitability of LAMP-OSD to various POC use scenarios, such as healthcare clinics and self-testing. Both assays performed accurately on the GeneTiger, a machine that automates both incubation and fluorogenic readout of LAMP-OSD assays, as well as the miniPCR machine with visual readout using an inexpensive handheld transilluminator. We could detect as few as eight synthetic MPXV DNA copies/reaction in only 30 min with simple visual readout of the tests by observing for presence (positive) or absence (negative) of bright green OSD fluorescence.

## METHODS

### Chemicals and reagents

Analytical grade chemicals were obtained from Sigma-Aldrich (St. Louis, MO, USA) unless otherwise indicated. Bst 2.0 DNA polymerase and isothermal amplification buffer were acquired from New England Biolabs (NEB, Ipswich, MA, USA). Gene blocks, LAMP primers, and OSD probe strands (**Table 1**) were purchased from Integrated DNA Technologies (IDT, Coralville, IA, USA).

**Table 1.**
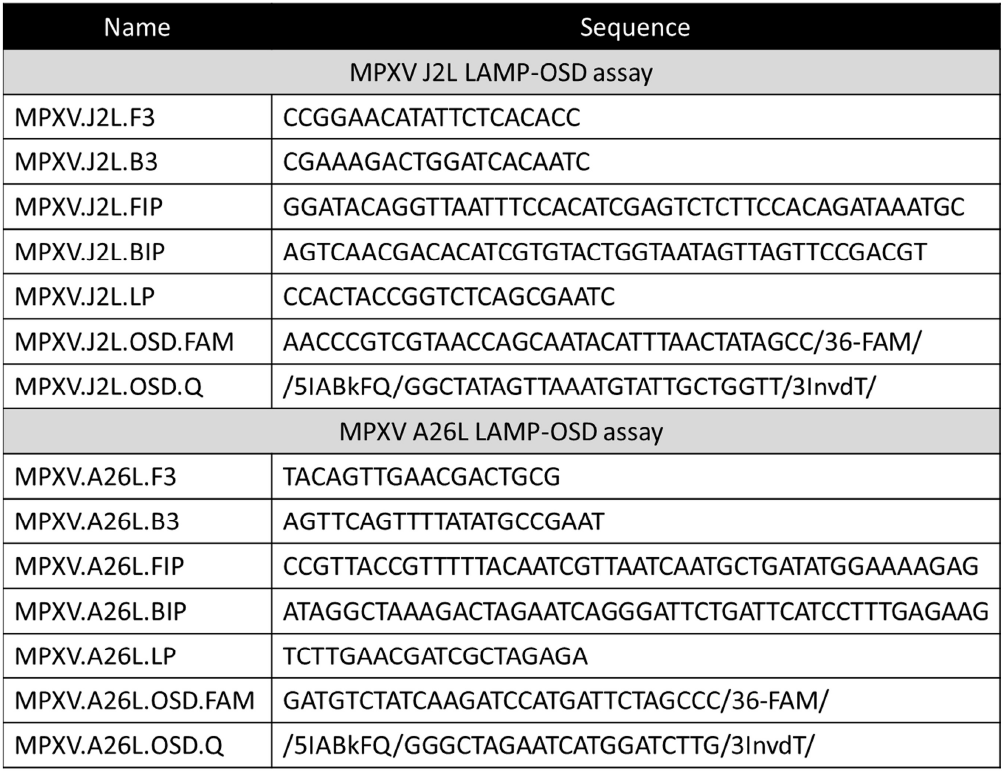
Primer and probe sequences.

### Primer and probe design

We designed two LAMP-OSD assays for clades 2 and 3 MPXV as follows. We built the MPXV J2L LAMP-OSD assay by designing a new LAMP primer set using the Primer Explorer v5 software (Eiken, Japan) (**Table 1**). These primers amplify a polymorphic portion of the MPXV J2L gene that includes a distinctive trinucleotide insertion/deletion (indel) found in clades 2 and 3 genomes and absent in clade 1 genomes (**Figure 2**). We designed the J2L LAMP primers such that this polymorphic locus is positioned towards one end of the loop sequence between the F1 and F2 regions of the amplicons (**Figure 2**). We engineered the J2L assay OSD probe, directed at this loop, to recognize this polymorphism via interaction with its toehold [22]. We derived the MPXV A26L LAMP-OSD assay primers using previously reported LAMP primers [6] that amplify a clade 2 and 3 specific region between MPXV A26L and A27L genes. We replaced the LF loop primer specific to the sequence between F1 and F2 primer specific sites with the OSD reporter.

**Figure 2.**
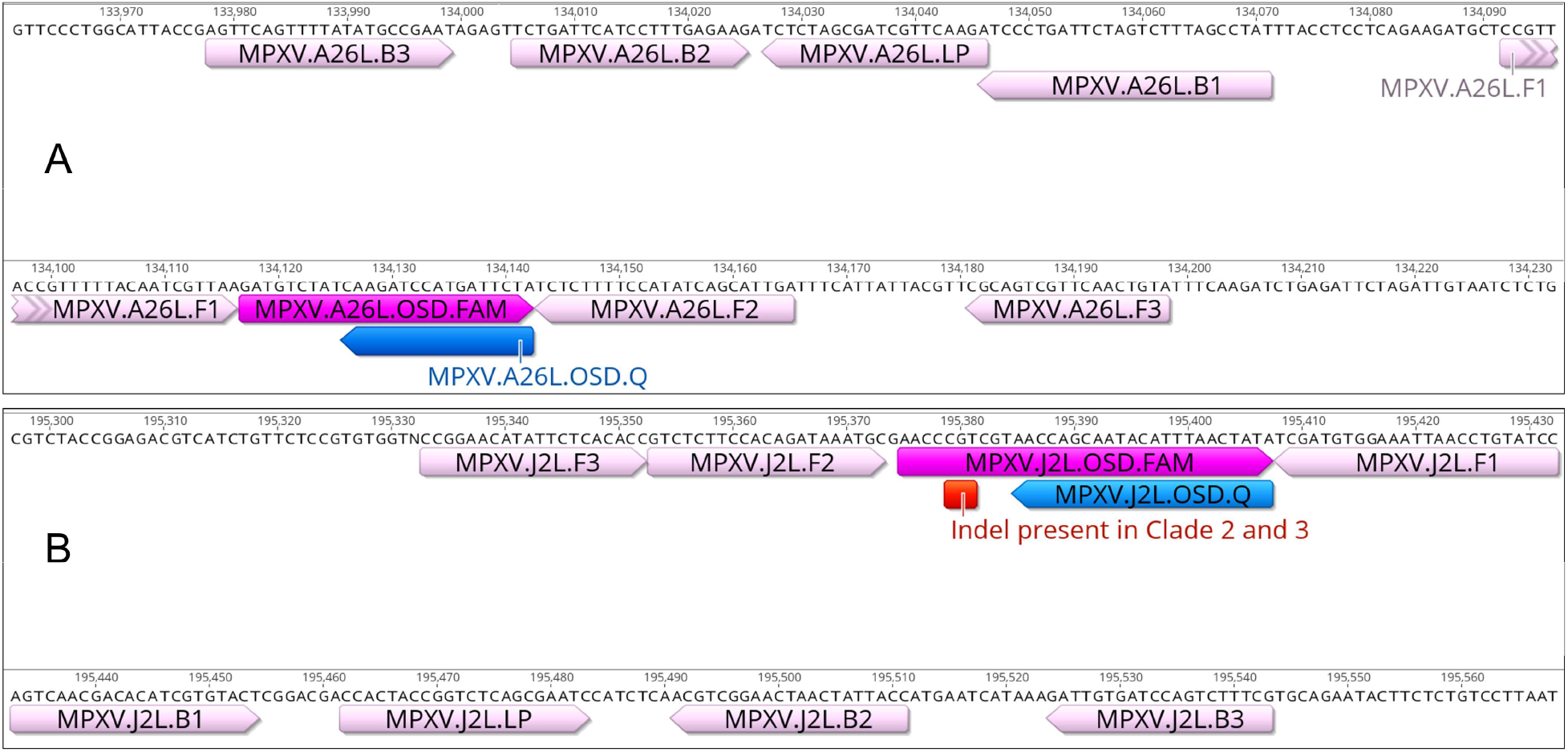
LAMP primer and OSD probe binding sequences in the MPXV genome (Monkeypox/PT0001/2022 [3]). Binding regions for A26L and J2L primers and OSD probes are depicted in panels A and B, respectively. Forward and reverse directions of the annotations correspond to the sense or antisense sequence. The fluorophore-labeled OSD strand is indicated as ‘OSD.FAM’ and the quencher-labeled OSD strand is indicated as ‘OSD.Q’. The trinucleotide indel present in clades 2 and 3 MPXV genomes is annotated with a red bar.

We designed both J2L and A26L LAMP assay OSD probes using our previously reported design rules and the NUPACK nucleic acid analysis software [22, 30, 31]. Briefly, we used a sequence between the F1 and F2 primer specific regions of the MPXV genome as the OSD long strand, which was labeled at its 3’-end with fluorescein (36-FAM). We designated a 10 (J2L OSD) or 9 (A26L OSD) nucleotide long section at the 5’-end of the long strand as the toehold. We designated the sequence reverse complementary to the remaining portion of long strand as the OSD short strand. We added three to four random basepairs at the 3’-end of the long strand and the 5’-end of the short strand to increase hemiduplex stability at LAMP amplification temperatures. The 5’-end of the short strand was labeled with an Iowa Black FQ (5IABkFQ) quencher moiety while the 3’-end was blocked against polymerase-mediated extension by appending an inverted dT nucleotide (3InvdT). We prepared hemiduplex OSD probes by mixing 1 µM of the long strand with 3 µM of the corresponding short strand in 1x isothermal buffer (20 mM Tris-HCl, 10 mM (NH_4_)_2_SO_4_, 50 mM KCl, 2 Mm MgSO_4_, 0.1% Tween® 20, pH 8.8@25°C). Annealing was performed by incubating this mixture at 95 °C for 1 min followed by slow cooling to 25 °C at the rate of 0.1 °C/sec.

### LAMP-OSD assay

We performed LAMP-OSD assays in 25 µL volume of 1x isothermal buffer containing 1 mM deoxyribonucleotides, 1.6 µM each of FIP and BIP, 0.8 µM of LP, 0.4 µM each of F3 and B3, 1M betaine, 3 mM additional MgSO_4_, 100 nM of fluorescein-labeled OSD long strand annealed with a three-fold excess of the quencher-labeled short strand, and 16 units of Bst 2.0 DNA polymerase. We used different amounts of synthetic double stranded DNA fragments (gBlocks™, IDT) as templates for amplification. Some negative control assays received 23 ng of human genomic DNA (Promega, Madison, WI) instead of synthetic MPXV templates. For real-time analysis of amplification kinetics, we assembled the LAMP-OSD reactions in a 96-well plate and incubated in a LightCycler 96 real-time PCR machine (Roche, Switzerland) programmed to hold the samples at 60 °C and measure OSD fluorescence at intervals of 3 mins. We generated amplification curves using the LightCycler 96 software. For assays operated on the GeneTiger instrument (GeneTiger, USA), we placed the LAMP-OSD assays in clear thin-walled 0.2 mL PCR tubes and loaded them on a GeneTiger rotary cartridge with positions marked 0 to 5 for accommodating 6 reaction tubes. Since we ran 5 LAMP-OSD reactions in an assay, we placed a tube containing 25 µL of water in position 5 to balance the cartridge. We placed the loaded cartridge in a GeneTiger programmed to hold the reactions at 60 °C for 60 min and capture images of OSD fluorescence in each tube at regular intervals. We noted the automated results at assay endpoint computed by the onboard GeneTiger neural net algorithms, from the instrument on-screen display and from the database available at https://genetiger.com/. We also imaged endpoint LAMP-OSD assay fluorescence using a cellphone camera by placing the GeneTiger cartridge with the assay tubes in a blue light transilluminator. For LAMP-OSD assays operated on a miniPCR machine (miniPCR Bio, Cambridge MA, USA), we placed the reactions in 0.2 mL thin-walled PCR tubes and placed them in a miniPCR set to hold the reactions at 60 °C for 30 min. At endpoint, we placed the tubes under a blue light transilluminator for visual readout of presence or absence of OSD fluorescence. We acquired images of assay tubes in the transilluminator using a cellphone camera.

## RESULTS

### Real-time amplification kinetics of MPXV LAMP-OSD assays

Assaying for two or more target-specific sequences often improves diagnostic accuracy [32, 33]. Therefore, we developed two LAMP-OSD assays, termed A26L and J2L assays, for rapid detection of clades 2 and 3 MPXV that include strains responsible for the current global monkeypox outbreak. The A26L LAMP-OSD assay uses five primers (FIP, BIP, F3, B3, and one LP) derived from a previously reported 6-primer LAMP assay specific to a region between A26L and A27L genes of clades 2 and 3 MPXV [6]. We replaced the second loop primer that bound the longer loop sequence of the amplicon with the A26L OSD probe. We have previously used this strategy to reduce false positive signal in several LAMP primer sets reported for the SARS-CoV-2 virus [23]. We designed the five primers for the J2L LAMP-OSD assay *de novo* to amplify a region of the J2L gene that includes a clade-specific indel. In particular, the sequence CGT found at nucleotide position 510-512 of the J2L coding sequence in clades 2 and 3 genomes is absent in clade 1 genomes (**Figure 2**). A TaqMan quantitative PCR assay for clades 2 and 3 MPXV also targeted this genomic region [34]. To detect this trinucleotide indel in J2L LAMP amplicons, and hence identify clades 2 and 3 MPXV, we constrained LAMP primer design to position the trinucleotide within the OSD toe-hold binding region of the amplicon loop sequence. It has been shown that toehold binding strength has significant impact on strand displacement kinetics [35]. OSD toeholds can be designed such that even single mismatches within the toehold abolish strand displacement and prevent signal [22].

To evaluate the amplification kinetics of the J2L and A26L LAMP-OSD assays, we seeded several replicate reactions with different amounts of synthetic MPXV double stranded DNA templates and incubated the assays at 60 °C for 60 min. We observed that OSD fluorescence increased over time in all reactions containing MPXV DNA templates while fluorescence in reactions lacking specific templates remained at baseline (**Figure 3**). These results indicate that both MPXV LAMP-OSD assays could readily detect their specific synthetic DNA templates and as few as eight copies of templates yielded a strong OSD signal above noise within 30 min.

**Figure 3.**
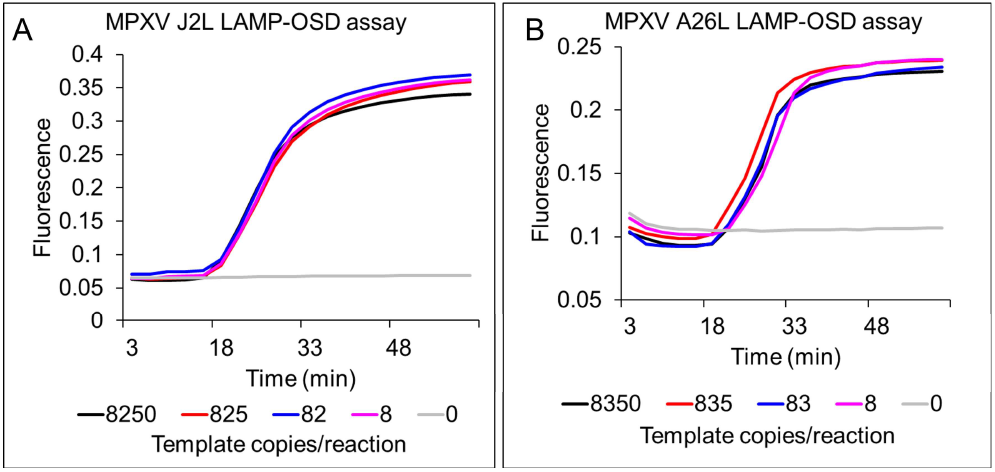
Real-time amplification kinetics of MPXV LAMP-OSD assays. OSD fluorescence values measured in real-time during LAMP amplification for J2L (panel A) and A26L (panel B) LAMP-OSD assays are depicted as amplification curves color-coded to indicate the number of synthetic MPXV double stranded DNA template copies per reaction. Data are representative of three biological replicates.

### Demonstration of the MPXV LAMP-OSD assays on portable devices

Having demonstrated the fast amplification kinetics and single digit template detection ability of both MPXV LAMP-OSD assays, we sought to demonstrate POC amenable implementation of the LAMP-OSD assays. Therefore, we tested assay performance on two different relatively inexpensive portable devices – the GeneTiger (cost: $1500) and the miniPCR16 (cost: $795). GeneTiger (https://genetiger.com/) has been customized for user friendly portable automation of fluorogenic LAMP-OSD assays with cartridge-based parallel operation of 6 independent assay tubes and both on-device and cloud-based reporting of analyzed test results. It is compact (13 cm x 19 cm x 8 cm; 1.5 kg) and can be operated for a full day on a single battery. The miniPCR16 (https://www.minipcr.com/) has 16-tube capacity and is compatible with Mac, PC, android, and iOS for controlling operation. It can be easily programmed for single temperature incubation of LAMP-OSD assays and yes/no results can be read at endpoint by visual inspection of presence or absence of bright OSD fluorescence using an inexpensive transilluminator.

To test performance of the MPXV LAMP-OSD assays on a GeneTiger, we assembled several replicate A26L and J2L reactions and challenged them with different amounts of synthetic MPXV DNA templates. We loaded these LAMP-OSD reaction tubes at positions 0 to 4 of a GeneTiger cartridge, such that reactions containing decreasing template amounts were located at positions 0 to 3 while the tube in position 4 lacked specific templates. Following 60 min of amplification at 60 °C the endpoint test results called by the GeneTiger indicated tubes 0, 1, 2, and 3 to be positive and tube 4 to be negative (**Figures 4A and 4D**).

**Figure 4.**
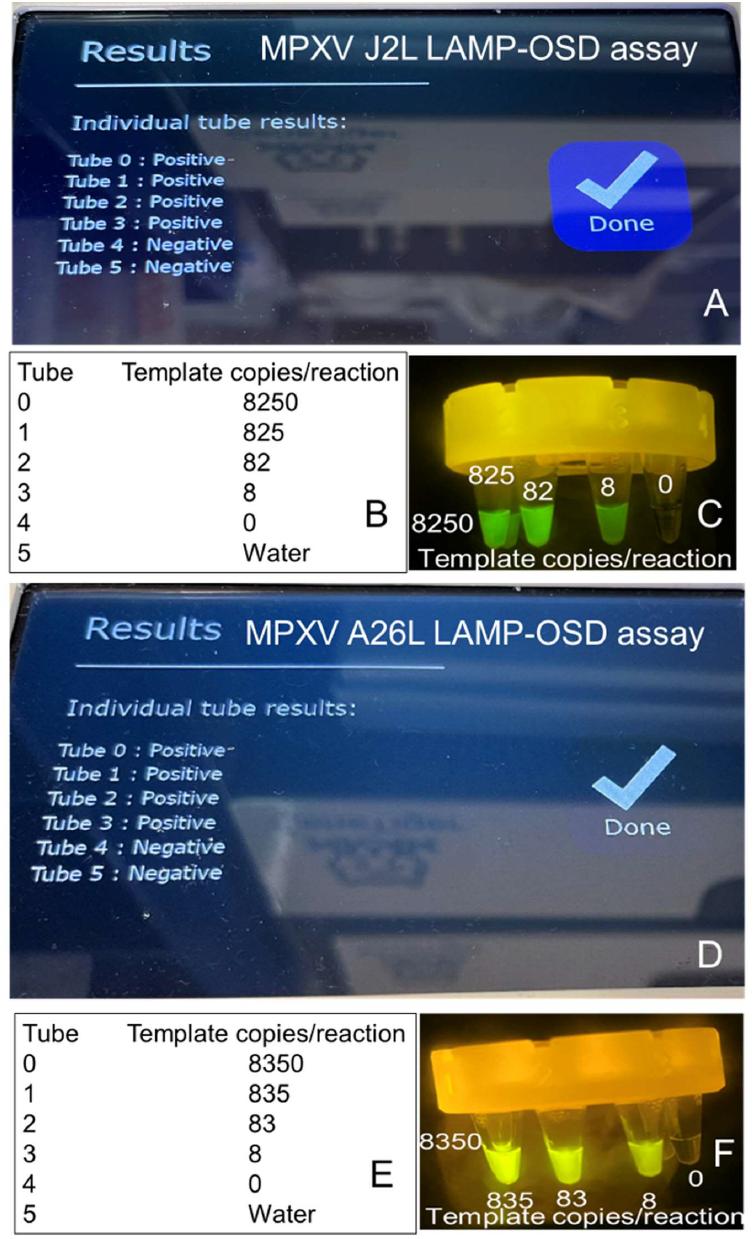
Performance of MPXV LAMP-OSD assays on a portable GeneTiger device. Images of GeneTiger screens displaying the automated results for each tube of J2L (panel A) or A26L (panel D) LAMP-OSD assay at endpoint are depicted. Number of synthetic MPXV double stranded DNA template copies/reaction tube is indicated in panels B (J2L assay) and E (A26L assay). Images of OSD fluorescence in J2L and A26L LAMP-OSD assay tubes in GeneTiger cartridges taken at reaction endpoint using a cellphone camera and a transilluminator are depicted in panels C and F. Data are representative of two biological replicates.

These automated results corresponded accurately with the presence or absence of MPXV DNA templates in the assay tubes (**Figures 4B and 4E**). Moreover, as few as eight MPXV DNA templates yielded a positive LAMP-OSD result on the GeneTiger. We also visually verified the accuracy of GeneTiger results by removing the cartridge from the machine and capturing an image of OSD fluorescence in the tubes using a cellphone camera and a blue light transilluminator. We observed bright green fluorescence in all LAMP-OSD reaction tubes that we had seeded with MPXV DNA templates (**Figures 4C and 4F**). Meanwhile, reaction tubes lacking specific templates were as dark as a tube containing water (position 5 on the GeneTiger cartridge). These results indicate that MPXV LAMP-OSD assays automated on the GeneTiger perform accurately.

We evaluated assay performance in a miniPCR16 device in a similar manner by testing amplification in several replicate A26L and J2L LAMP-OSD assays containing either no templates, human genomic DNA, or different amounts of synthetic MPXV DNA templates. Following 30 min of amplification in the miniPCR, when we placed the assay tubes in a blue light transilluminator, we observed bright green fluorescence in all assay tubes containing MPXV specific DNA templates (**Figure 5**). Meanwhile, tubes lacking any templates or containing human genomic DNA as templates remained dark (**Figure 5**). These results indicate that LAMP-OSD assays perform accurately in a miniPCR machine and can be easily read visually using minimal equipment. Overall, these results signify the robustness of LAMP-OSD, which enables its facile compatibility with low resource operation suitable for field-based, POC, and even home-based use for actionable diagnostics.

**Figure 5.**
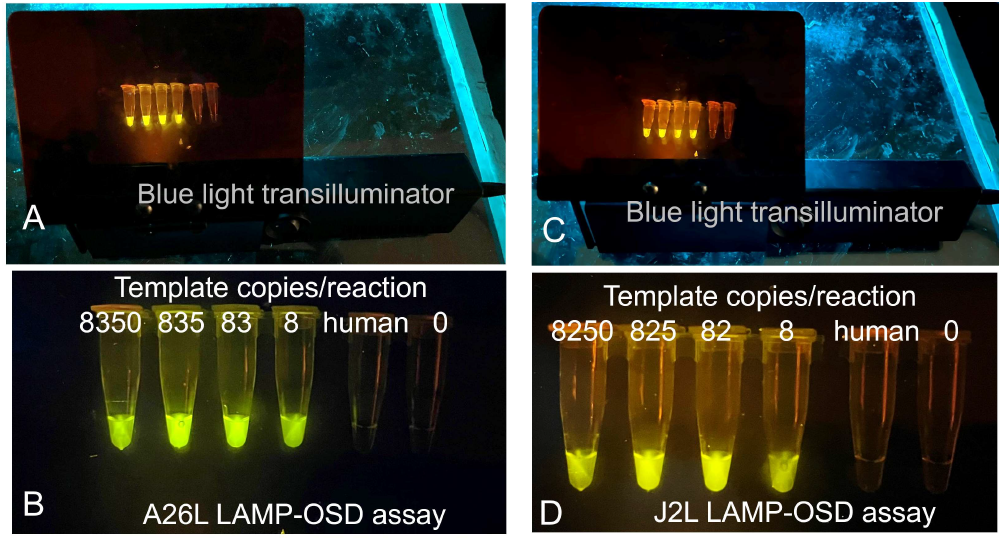
Performance of MPXV LAMP-OSD assays using a miniPCR16 machine. Images of endpoint OSD fluorescence in A26L (panels A and B) and J2L (panels C and D) LAMP-OSD assays performed in a miniPCR16 machine are depicted. Panels A and C are zoomed out images of the assay tubes in a transilluminator while panels B and D show close-up views of the same assay tubes. Number of synthetic MPXV double stranded DNA template copies/reaction tube is indicated in panels B (A26L assay) and D (J2L assay). Data are representative of two biological replicates.

## DISCUSSION

We have demonstrated two LAMP-OSD assays for detection of clades 2 and 3 MPXV (also known as the West African MPXV) that include the current outbreak strains of MPXV [36]. Both assays can readily detect single digit copies of their target DNA within 30 min. Furthermore, we demonstrated POC amenable operation of both assays using two portable devices - the GeneTiger that automated both assay incubation and readout and the miniPCR that held the assays at the amplification temperature while endpoint readout was obtained visually using a trans-illuminator. We envision that a real-world implementation of this diagnostic platform would require only four user-mediated steps (**Figure 6**). In Step 1, the specimens, such as skin lesion samples, would be processed by a short incubation at 95 °C to both inactivate the virions and extract viral nucleic acids. MPXV is not heat stable and previous reports have demonstrated the feasibility of detecting Orthopoxvirus DNA by direct PCR analysis of heat-treated human and animal specimens without needing prior DNA purification [37, 38]. Furthermore, compatibility of LAMP-OSD reactions with direct analysis of crude biological specimens, such as crushed insects, human saliva, and environmental water, is well-documented [24, 26]. Step 2 would involve transfer of an aliquot of the heated sample into ready-to-use MPXV LAMP-OSD assays. Freeze-dried LAMP-OSD reactions are stable at ambient temperature for at least several months and can be used for diagnostic tests simply by rehydration with assay buffer and test specimens [28]. In Step 3, the user will load the assays onto a portable device to initiate assay incubation, and in the last step, depending on the device, presence or absence of OSD fluorescence, which is indicative of presence or absence of target nucleic acids, will be recorded automatically or manually by visual inspection. Overall, these Steps are highly amenable to assays with off-the-shelf devices, such as the low-cost and portable GeneTiger or miniPCR, and could be readily packaged into a single device devoted to the assay.

**Figure 6.**
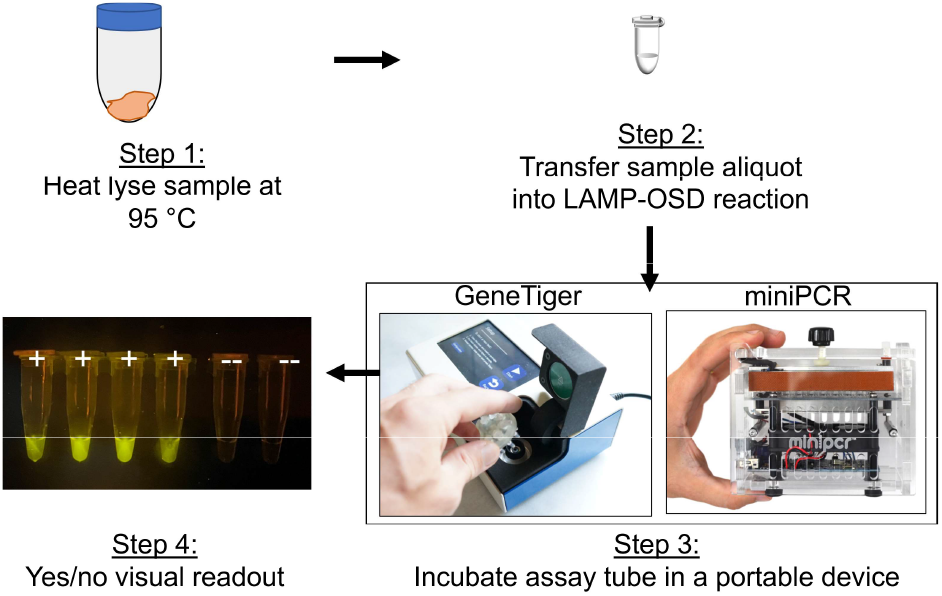
Proposed steps for implementing MPXV POC diagnostics using LAMP-OSD assays.

As we have previously shown for numerous other diseases and biomedical indications, rapid, sequence-specific readout mediated via strand displacement makes LAMP-OSD assays extremely suitable for point-of-need detection. This is especially true in the current MPXV outbreak, given the need to enable personal testing options that can run in parallel to the public health infrastructure. As MPXV outbreak strains continue to evolve, diagnostic surety may be enhanced by composing assays into multiplex reactions where OSDs can be used to logically compute the presence or absence of multiple viral amplicons [23, 26], thereby allowing the same platform to evolve along with the virus through simple changes in chemistry.

## Data Availability

All data in the present work and contained in the manuscript.

## ACKNOWLEDGEMENTS

This work was supported by the Welch Foundation (F-1654) and the National Institutes of Health (1R01EB027202-01A1 and 3R01EB027202-01A1S1).

## REFERENCES

1. WHO. Monkeypox. 2022. Available from: https://www.who.int/news-room/fact-sheets/detail/monkeypox.

2. Happi C, Adetifa I, Mbala P, Njouom R, Nakoune E, Happi A, et al. Urgent need for a non-discriminatory and non-stigmatizing nomenclature for monkeypox virus. 2022. Available from: https://virological.org/t/urgent-need-for-a-non-discriminatory-and-non-stigmatizing-nomenclature-for-monkeypox-virus/853.

3. Isidro J, Borges V, Pinto M, Sobral D, Santos JD, Nunes A, et al. Phylogenomic characterization and signs of microevolution in the 2022 multi-country outbreak of monkeypox virus. Nature medicine. 2022. Epub 20220624. doi: 10.1038/s41591-022-01907-y. PubMed PMID: 35750157.

4. CDC. 2022 U.S. Monkeypox Outbreak 2022. Available from: https://www.cdc.gov/poxvirus/monkeypox/response/2022/index.html.

5. Titanji BK, Tegomoh B, Nematollahi S, Konomos M, Kulkarni PA. Monkeypox: A Contemporary Review for Healthcare Professionals. Open Forum Infect Dis. 2022;9(7):ofac310. Epub 20220623. doi: 10.1093/ofid/ofac310. PubMed PMID: 35891689; PubMed Central PMCID: PMCPMC9307103.

6. Iizuka I, Saijo M, Shiota T, Ami Y, Suzaki Y, Nagata N, et al. Loop-mediated isothermal amplification-based diagnostic assay for monkeypox virus infections. J Med Virol. 2009;81(6):1102–8. doi: 10.1002/jmv.21494. PubMed PMID: 19382264.

7. Davi SD, Kissenkötter J, Faye M, Böhlken-Fascher S, Stahl-Hennig C, Faye O, et al. Recombinase polymerase amplification assay for rapid detection of Monkeypox virus. Diagn Microbiol Infect Dis. 2019;95(1):41–5. Epub 20190411. doi: 10.1016/j.diagmicrobio.2019.03.015. PubMed PMID: 31126795.

8. Bhadra S, Nguyen V, Torres JA, Kar S, Fadanka S, Gandini C, et al. Producing molecular biology reagents without purification. Plos One. 2021;16(6):e0252507. Epub 2021/06/02. doi: 10.1371/journal.pone.0252507. PubMed PMID: 34061896; PubMed Central PMCID: PMCPMC8168896 SF is Executive Director of Mboalab, a Cameroonian registered company. JM is an unpaid Executive Director of Beneficial Bio Ltd, a UK not-for-profit company limited by guarantee and a volunteer board member of the Gathering for Open Science Hardware Inc, a US 501c3 nonprofit. ADE and SB are named inventors on pending US patent application S20200399679A1 covering cellular reagents, assigned to the University of Texas System. These competing interests do not alter our adherence to PLOS ONE policies on sharing data and materials.

9. Bhadra S, Pothukuchy A, Shroff R, Cole AW, Byrom M, Ellefson JW, et al. Cellular reagents for diagnostics and synthetic biology. Plos One. 2018;13(8):e0201681. Epub 2018/08/16. doi: 10.1371/journal.pone.0201681. PubMed PMID: 30110361; PubMed Central PMCID: PMCPMC6093680.

10. Bhadra S, Paik I, Torres J-A, Fadanka S, Gandini C, Akligoh H, et al. Preparation and Use of Cellular Reagents: A Low-resource Molecular Biology Reagent Platform. Current Protocols. 2022;2(3):e387. doi: https://doi.org/10.1002/cpz1.387.

11. Paik I, Bhadra S, Ellington AD. Charge Engineering Improves the Performance of Bst DNA Polymerase Fusions. ACS Synth Biol. 2022;11(4):1488–96. Epub 20220323. doi: 10.1021/acssynbio.1c00559. PubMed PMID: 35320674.

12. Paik I, Ngo PHT, Shroff R, Diaz DJ, Maranhao AC, Walker DJF, et al. Improved Bst DNA Polymerase Variants Derived via a Machine Learning Approach. Biochemistry. 2021. Epub 20211111. doi: 10.1021/acs.biochem.1c00451. PubMed PMID: 34762799.

13. Notomi T, Okayama H, Masubuchi H, Yonekawa T, Watanabe K, Amino N, et al. Loop-mediated isothermal amplification of DNA. Nucleic Acids Res. 2000;28(12):E63. Epub 2000/06/28. PubMed PMID: 10871386; PubMed Central PMCID: PMCPMC102748.

14. Nagamine K, Hase T, Notomi T. Accelerated reaction by loop-mediated isothermal amplification using loop primers. Mol Cell Probes. 2002;16(3):223–9. Epub 2002/07/30. PubMed PMID: 12144774.

15. Gandelman O, Jackson R, Kiddle G, Tisi L. Loop-mediated amplification accelerated by stem primers. Int J Mol Sci. 2011;12(12):9108–24. Epub 2012/01/25. doi: 10.3390/ijms12129108. PubMed PMID: 22272122; PubMed Central PMCID: PMC3257119.

16. Martineau RL, Murray SA, Ci S, Gao W, Chao SH, Meldrum DR. Improved Performance of Loop-Mediated Isothermal Amplification Assays via Swarm Priming. Anal Chem. 2017;89(1):625–32. Epub 20161220. doi: 10.1021/acs.analchem.6b02578. PubMed PMID: 27809497.

17. Mori Y, Nagamine K, Tomita N, Notomi T. Detection of loop-mediated isothermal amplification reaction by turbidity derived from magnesium pyrophosphate formation. Biochem Biophys Res Commun. 2001;289(1):150–4. doi: 10.1006/bbrc.2001.5921. PubMed PMID: 11708792.

18. Oscorbin IP, Belousova EA, Zakabunin AI, Boyarskikh UA, Filipenko ML. Comparison of fluorescent intercalating dyes for quantitative loop-mediated isothermal amplification (qLAMP). Biotechniques. 2016;61(1):20–5. Epub 20160701. doi: 10.2144/000114432. PubMed PMID: 27401670.

19. Tanner NA, Zhang YH, Evans TC. Visual detection of isothermal nucleic acid amplification using pH-sensitive dyes. Biotechniques. 2015;58(2):59-68. 10.2144/000114253. PubMed WOS:000349423400003. doi: PMID:

20. Zhang DY, Seelig G. Dynamic DNA nanotechnology using strand-displacement reactions. Nat Chem. 2011;3(2):103–13. doi: Doi 10.1038/Nchem.957. PubMed PMID: ISI:000286505700006.

21. Holland PM, Abramson RD, Watson R, Gelfand DH. Detection of specific polymerase chain reaction product by utilizing the 5’ 3’ exonuclease activity of Thermus aquaticus DNA polymerase. Proceedings of the National Academy of Sciences of the United States of America. 1991;88(16):7276–80. doi: 10.1073/pnas.88.16.7276. PubMed PMID: 1871133; PubMed Central PMCID: PMCPMC52277.

22. Jiang YS, Bhadra S, Li B, Wu YR, Milligan JN, Ellington AD. Robust strand exchange reactions for the sequence-specific, real-time detection of nucleic acid amplicons. Anal Chem. 2015;87(6):3314–20. Epub 2015/02/25. doi: 10.1021/ac504387c. PubMed PMID: 25708458.

23. Bhadra S, Riedel TE, Lakhotia S, Tran ND, Ellington AD. High-Surety Isothermal Amplification and Detection of SARS-CoV-2. mSphere. 2021;6(3):e00911–20. doi: 10.1128/mSphere.00911-20.

24. Bhadra S, Riedel TE, Saldana MA, Hegde S, Pederson N, Hughes GL, et al. Direct nucleic acid analysis of mosquitoes for high fidelity species identification and detection of Wolbachia using a cellphone. PLoS Negl Trop Dis. 2018;12(8):e0006671. Epub 2018/08/31. doi: 10.1371/journal.pntd.0006671. PubMed PMID: 30161131; PubMed Central PMCID: PMCPMC6116922.

25. Bhadra S, Jiang YS, Kumar MR, Johnson RF, Hensley LE, Ellington AD. Real-time sequence-validated loop-mediated isothermal amplification assays for detection of Middle East respiratory syndrome coronavirus (MERS-CoV). Plos One. 2015;10(4):e0123126. Epub 2015/04/10. doi: 10.1371/journal.pone.0123126. PubMed PMID: 25856093; PubMed Central PMCID: PMCPMC4391951.

26. Bhadra S, Saldana MA, Han HG, Hughes GL, Ellington AD. Simultaneous Detection of Different Zika Virus Lineages via Molecular Computation in a Point-of-Care Assay. Viruses. 2018;10(12):Preprint accessible at: doi: https://doi.org/10.1101/424440. Epub 2018/12/19. doi: 10.3390/v10120714. PubMed PMID: 30558136; PubMed Central PMCID: PMCPMC6316447.

27. Du Y, Hughes RA, Bhadra S, Jiang YS, Ellington AD, Li B. A Sweet Spot for Molecular Diagnostics: Coupling Isothermal Amplification and Strand Exchange Circuits to Glucometers. Sci Rep. 2015;5:11039. Epub 2015/06/09. doi: 10.1038/srep11039. PubMed PMID: 26050646; PubMed Central PMCID: PMCPMC4458886.

28. Jiang YS, Riedel TE, Popoola JA, Morrow BR, Cai S, Ellington AD, et al. Portable platform for rapid in-field identification of human fecal pollution in water. Water Res. 2018;131:186–95. Epub 2017/12/27. doi: 10.1016/j.watres.2017.12.023. PubMed PMID: 29278789; PubMed Central PMCID: PMCPMC5999531.

29. Jiang YS, Stacy A, Whiteley M, Ellington AD, Bhadra S. Amplicon Competition Enables End-Point Quantitation of Nucleic Acids Following Isothermal Amplification. Chembiochem. 2017;18(17):1692–5. Epub 2017/06/20. doi: 10.1002/cbic.201700317. PubMed PMID: 28628741; PubMed Central PMCID: PMCPMC5890436.

30. Zadeh JN, Steenberg CD, Bois JS, Wolfe BR, Pierce MB, Khan AR, et al. NUPACK: Analysis and design of nucleic acid systems. Journal of computational chemistry. 2011;32(1):170–3. Epub 2010/07/21. doi: 10.1002/jcc.21596. PubMed PMID: 20645303.

31. Zadeh JN, Steenberg CD, Bois JS, Wolfe BR, Pierce MB, Khan AR, et al. NUPACK: Analysis and design of nucleic acid systems. J Comput Chem. 2011;32(1):170–3. Epub 2010/07/21. doi: 10.1002/jcc.21596. PubMed PMID: 20645303.

32. Molina F, Lopez-Acedo E, Tabla R, Roa I, Gomez A, Rebollo JE. Improved detection of Escherichia coli and coliform bacteria by multiplex PCR. BMC Biotechnol. 2015;15:48. Epub 2015/06/05. doi: 10.1186/s12896-015-0168-2. PubMed PMID: 26040540; PubMed Central PMCID: PMCPMC4453288.

33. Riojas MA, Kiss K, McKee ML, Hazbon MH. Multiplex PCR for species-level identification of Bacillus anthracis and detection of pXO1, pXO2, and related plasmids. Health Secur. 2015;13(2):122–9. Epub 2015/03/31. doi: 10.1089/hs.2014.0056. PubMed PMID: 25813976; PubMed Central PMCID: PMCPMC4394167.

34. Li Y, Zhao H, Wilkins K, Hughes C, Damon IK. Real-time PCR assays for the specific detection of monkeypox virus West African and Congo Basin strain DNA. Journal of Virological Methods. 2010;169(1):223–7. doi: https://doi.org/10.1016/j.jviromet.2010.07.012.

35. Zhang DY, Winfree E. Control of DNA strand displacement kinetics using toehold exchange. J Am Chem Soc. 2009;131(47):17303–14. Epub 2009/11/10. doi: 10.1021/ja906987s. PubMed PMID: 19894722.

36. Gigante CM, Korber B, Seabolt MH, Wilkins K, Davidson W, Rao AK, et al. Multiple lineages of Monkeypox virus detected in the United States, 2021-2022. bioRxiv. 2022:2022.06.10.495526. doi: 10.1101/2022.06.10.495526.

37. Abrahao JS, Drumond BP, Trindade Gde S, da Silva-Fernandes AT, Ferreira JM, Alves PA, et al. Rapid detection of Orthopoxvirus by semi-nested PCR directly from clinical specimens: a useful alternative for routine laboratories. J Med Virol. 2010;82(4):692–9. doi: 10.1002/jmv.21617. PubMed PMID: 20166167.

38. Rouhandeh H, Engler R, Taher M, Fouad A, Sells LL. Properties of monkey pox virus. Arch Gesamte Virusforsch. 1967;20(3):363–73. doi: 10.1007/BF01241954.

